# Role of anti-polyethylene glycol (PEG) antibodies in the allergic reactions and immunogenicity of PEG-containing Covid-19 vaccines

**DOI:** 10.1101/2022.10.03.22280227

**Authors:** Gergely Tibor Kozma, Tamás Mészáros, Petra Berényi, Réka Facskó, Zsófia Patkó, Csaba Zs. Oláh, Adrienne Nagy, Tamás Gyula Fülöp, Kathryn Anne Glatter, Tamás Radovits, Béla Merkely, János Szebeni

## Abstract

The polyethylene-glycol (PEG)-containing Covid-19 vaccines can cause hypersensitivity reactions (HSRs), or rarely, life-threatening anaphylaxis. A causal role of anti-PEG antibodies (Abs) has been proposed, but not yet proven in humans. The 191 blood donors in this study included 10 women and 5 men who displayed HSRs to Comirnaty or Spikevax Covid-19 vaccines with 3 anaphylaxis. 118 donors had pre-vaccination anti-PEG IgG/IgM values as measured by ELISA, of which >98% were over background regardless of age, indicating the presence of these Abs in almost everyone. Their values varied over 2-3 orders of magnitude and displayed strong left-skewed distribution with 3-4% of subjects having >15-30-fold higher values than the respective median. First, or booster injections with both vaccines led to significant rises of anti-PEG IgG/IgM with >10-fold rises in about ∼10% of Comirnaty, and all Spikevax recipients, measured at different times after the injections. The anti-PEG Ab levels measured within 4-months after the HSRs were significantly higher than those in nonreactors. Serial testing of plasma (n=361 tests) showed the SARS-CoV-2 neutralization IgG to vary over a broad range, with a trend for biphasic dose dependence on anti-PEG Abs. The highest prevalence of anti-PEG Ab positivity in human blood reported to date represents new information which can most easily be rationalized by daily exposure to common PEG-containing medications and/or household items. The significantly higher, HSR-non-coincidental blood level of anti-PEG Abs in hypersensitivity reactor vs. non-reactors, taken together with relevant clinical and experimental data in the literature, suggest that anti-PEG Ab supercarrier people might be at increased risk for HSRs to PEG-containing vaccines, which themselves can induce these Abs via bystander immunogenicity. Our data also raise the possibility that anti-PEG Abs might also contribute to the reduction of these vaccines’ virus neutralization efficacy. Thus, screening for anti-PEG Ab supercarriers may identify people at risk for HSRs or reduced vaccine effectiveness.

## INTRODUCTION

The efficacy of mRNA-lipid nanoparticle (LNP)-based Covid-19 vaccines (Comirnaty and Spikevax) in reducing death or severe illness from SARS-CoV-2 infections is well recognized. However, as with all vaccinations, these vaccines may also have side effects in some people. One of them is an allergic reaction, also known as a hypersensitivity reaction (HSR), whose most severe manifestation is anaphylaxis.

Anaphylactic reactions to these vaccines, which are behind the mandated post-vaccination observation of vaccinees, are very rare, and mostly controllable with epinephrine. Yet, there are still life-threatening reactions and the phenomenon still represents a problem for severely allergic people.^1-18^ Additionally, the specter of an allergic reaction helps fuel vaccine hesitancy.

Since the mRNA-LNP vaccines include polyethylene glycol (PEG) as an excipient, allergy to PEG has been proposed as the mechanism of anaphylaxis. However, over time, it has been shown that the overwhelming majority of these reactions are not IgE-mediated classic type I allergies against PEG,^1-17^ leaving the underlying cause unclear. Among the studies addressing this puzzle, the possible role of anti-PEG antibodies (Abs) was raised,^1, 17, 19, 20^ but conclusive experimental or clinical evidence was not presented to date.

Most recently, two studies reported the rise of anti-PEG Abs in Spikevax-vaccinated people,^21, 22^ implying specific binding of these Abs to PEG in the vaccine. In earlier studies, anti-PEG Abs were shown to cause rapid loss of efficacy along with increased the risk of HSRs to PEGylated urate oxidase,^23, 24^ and in another example, 3 anaphylactic reactions in patients having very high pre-existing anti-PEG Abs led to early termination of a Phase II clinical trial with the PEGylated RNA aptamer, Pegnivacogin.^25, 26^ These clinical data are consistent with animal studies showing anti-PEG Ab-mediated anaphylaxis^27^ and accelerated blood clearance (i.e., loss of efficacy) after i.v. injection of PEGylated liposomes.^28, 29^

Considering the parallelisms between the structures of mRNA-LNP vaccines and the above reactogenic nanomedicines with potential waning of efficacy, we hypothesized that similarly to these clinical and experimental examples, a contributing factor to the occasional anaphylactic reactivity of mRNA-LNP vaccines and the individual variation of antiviral Ab levels they induce could be associated with high levels of anti-PEG Abs.

Accordingly, we measured the plasma levels of anti-PEG IgG and IgM in allergic reactors and nonreactors to Comirnaty and Spikevax vaccinations, using non-PEGylated Covid-19 vaccines as controls. These anti-PEG Abs were also correlated with the levels of SARS-CoV-2 neutralizing Abs, taken as endpoint for vaccine efficacy, and relating the postvaccination anti-PEG Ab levels to the pre-vaccination values allowed us to investigate “bystander” anti-PEG immunogenicity, similar to that reported for Spikevax.^21, 22^

## RESULTS AND DISCUSSION

### Plasma anti-PEG IgG and IgM levels before vaccination: impacts of age, gender and cosmetic use

Fig. 1 shows the anti-PEG IgG (A) and IgM (B) levels and distribution in the blood of Covid-19 unvaccinated participants, and Table 1 lists the main statistical parameters derived from these data. Contrary to the 10-76% range of anti-PEG Ab positivity in various studies,^21, 25, 26, 30-34^ we found detectable levels of these Abs in 98-99% blood donors. Their distribution was highly left-skewed, with considerable, 2-3 orders of magnitude span between the lowest and highest values. The low, near baseline Ab levels in most samples is most easily rationalized by low level immunization via the skin or per os by daily exposure to PEG, an excipient in many oral, topical, and parenteral pharmaceutical products, hygiene and cosmetic items, and processed food and beverages.^35^ It is also important to mention that Abs against PEG and polysorbate 80 (PS-80), which is a branched PEG derivative, can mutually cross-react,^36-38^ and PS-80 is also widespread as an ingredient in drugs, vaccines, vitamins, food and drinks.^35, 39^ Thus, the anti-PEG ELISA does not necessarily distinguishes between these Abs.

**Figure 1.**
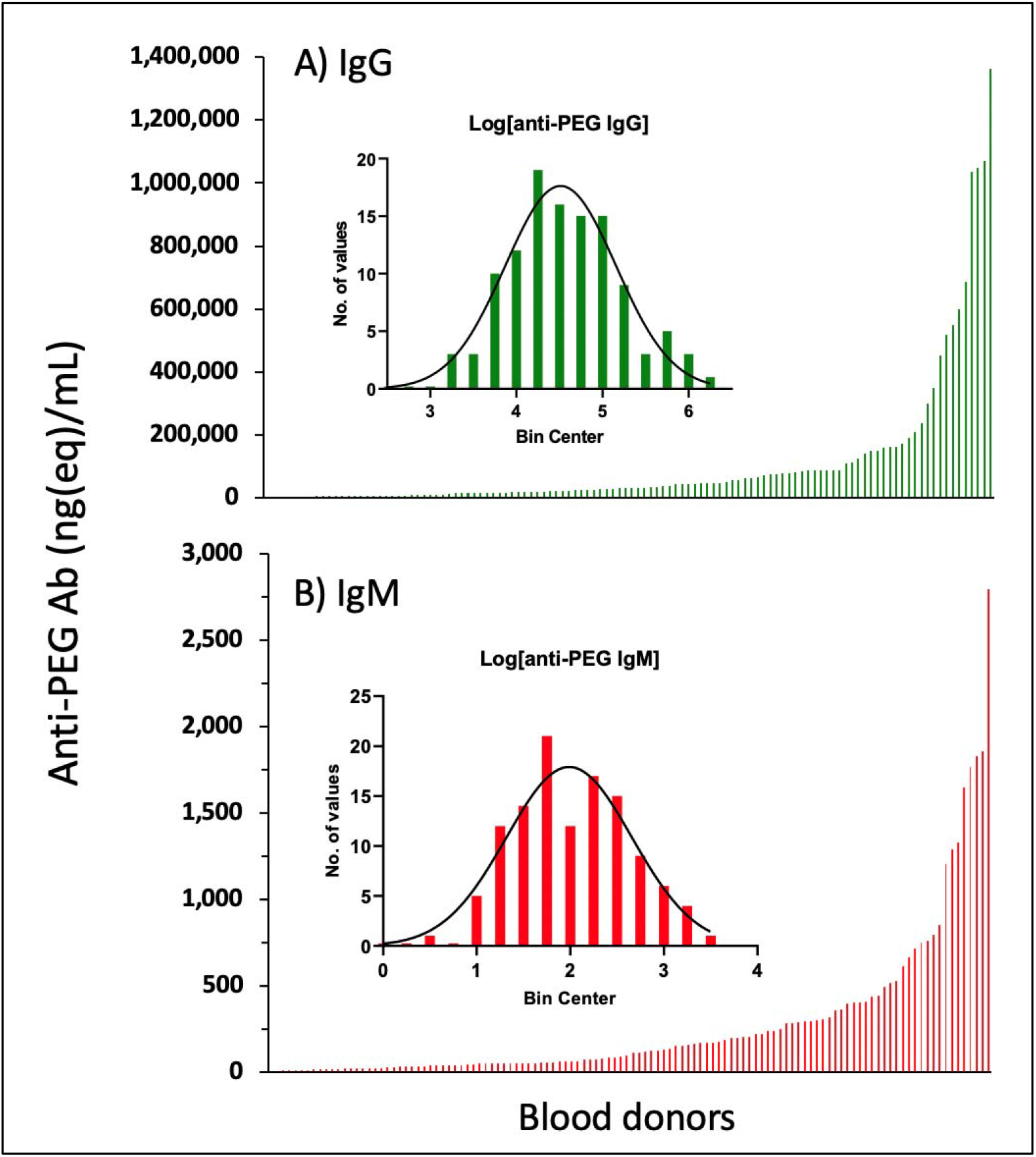
**Pre-vaccination anti-PEG Antibody Concentrations in a Mixed Population of Blood Donors. Anti-PEG-IgG (A, green) and anti-PEG-IgM (B, red) values were sorted in growing order. The bottom bars (which look like lines) are absolute Ab concentrations. The inserted probability distribution histograms are made from the log of absolute Ab levels, grouped into bins on the abscissa.^40^ The normality of distribution was established by the Shapiro-Wilk test. The anti-PEG Ab levels were determined as described in the Methods, and the descriptive statistics of these data are shown in Table 1.**

**Table 1.**
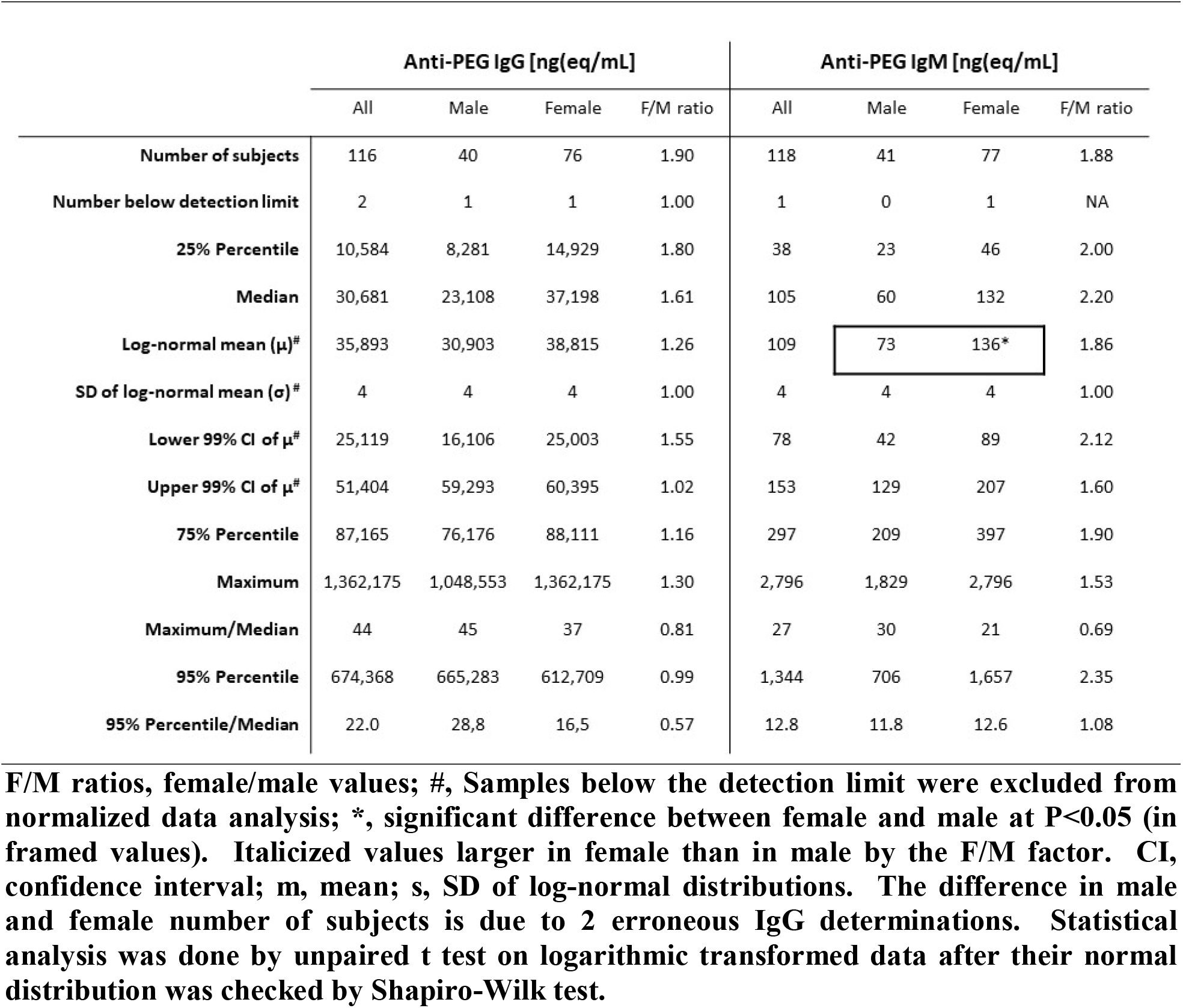
Descriptive statistics of pre-vaccination anti-PEG-IgG and IgM levels and distributions in humans.

Table 1 also shows that the prevalence and absolute levels of pre-vaccination anti-PEG IgM was significantly higher in females compared to males. Questioning about cosmetic use revealed that 1/3 of man were frequent cosmetic users versus >3/4 of women (Fig. 2A), and there was a trend for higher IgG (Fig 2B), and significantly higher anti-PEG IgM in women (Fig 2C).

**Figure 2.**
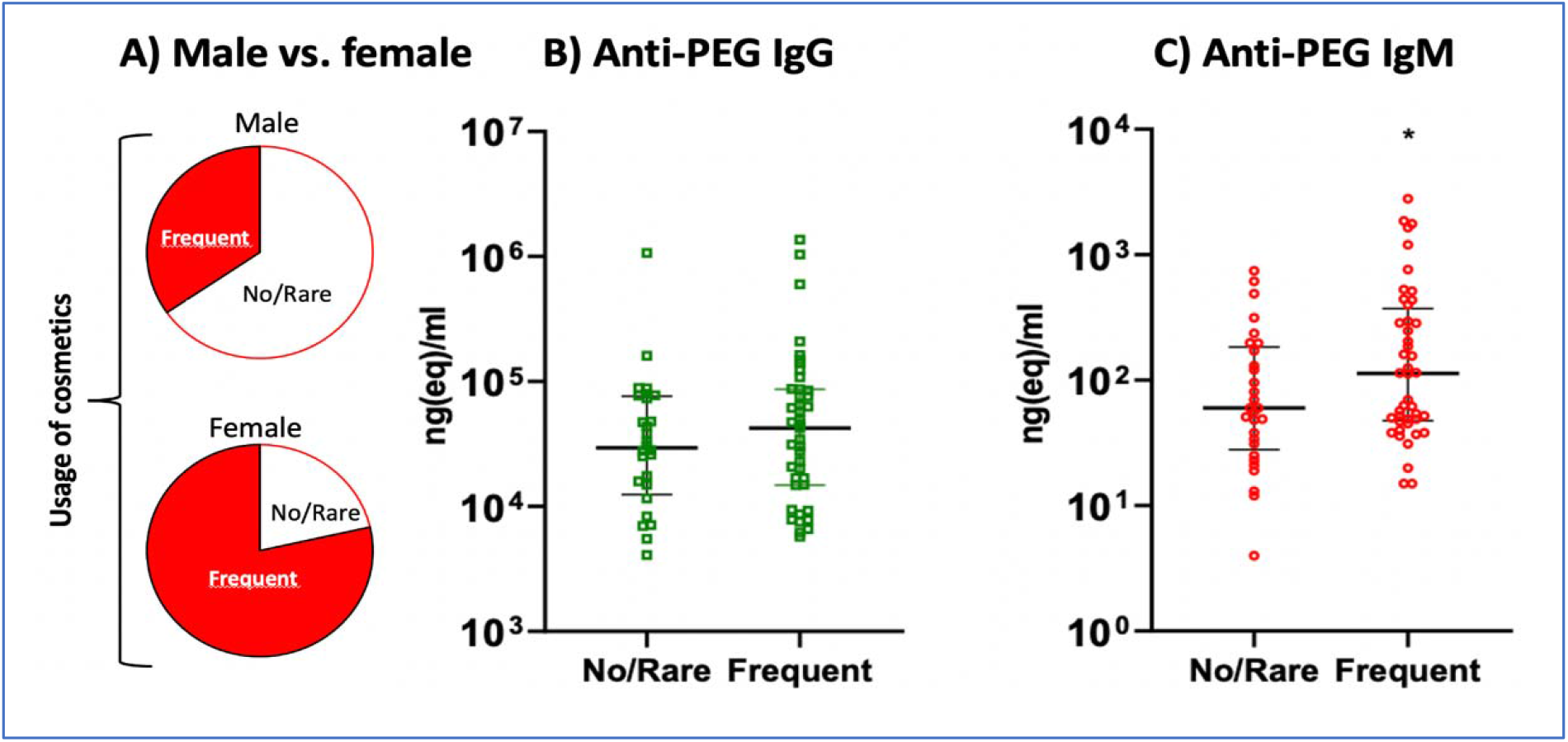
**Use of cosmetics by women and men and blood anti-PEG antibody levels in the two genders. A) Ratio of frequent cosmetic users vs. no users or rare users in men and women. B) Blood levels of anti-PEG-IgG (B) and anti-PEG-IgM (C) in frequent vs. no/rare cosmetic users before COVID-19 vaccination. The anti-PEG Ab levels were determined as described in the Methods. *P< 0.05 by 2-tailed t-test of logarithmic transformed data.**

Correlating the anti-PEG IgG or IgM levels with age showed no significant correlation (data not shown), and surprisingly, we found relatively high anti-PEG IgG (36 µg(eq)/mL) and IgM (401 ng(eq)/mL) even in a baby less than one year old. Eight children in the 6-11 year-old range also displayed high values (median of anti-PEG IgG and IgM were 27 µg(eq)/mL and 56 ng(eq)/mL, respectively).

We have also analyzed the relative changes of anti-PEG-IgG and IgM levels in the blood of 6 unvaccinated people in whom 2 measurements were available with different time intervals. There was substantial variation of anti-PEG IgM over time, while the anti-PEG IgG remained relatively constant in the same individuals (Supplement Fig S1-A). Although the n is too small for making conclusions from these data, they raise the possibility of differential IgM and IgG responses to transient PEG exposures that could take place in-between the blood withdrawals. Accordingly, there was no significant correlation between the anti-PEG IgM and IgG levels before vaccination (Fig. 1S-B). Nevertheless, we observed in 4 participants with a history of allergy (red squares) and 3 with mastocytosis (green triangles) maximal anti-PEG-IgM levels together with minimal anti-PEG IgG, or the opposite (Fig. 1S-B). These may be reflections of immune abnormalities in these subjects calling for further attention to the cause and consequences of the phenomenon. Our data also suggested higher anti-PEG IgG levels in those donors who suffered from persistent allergies due to dust mite, mold, cat hair, foods, numerous pollen type, chemicals, drugs, or cosmetics (data not shown).

Commenting on the higher prevalence of anti-PEG Ab positivity in our study relative to the literature,^21, 25, 26, 30-34^ the exact reason is not clear. The discrepancy may partly be due to the diverse quantitation methods for anti-PEG Abs in these studies,^36^ and partly to the different inclusion criteria of blood donors. For example, each referred reports on the prevalence of anti-PEG Abs in the healthy population or drug-treated patients^21, 25, 26, 30-34^ used different antigens and/or differing test conditions in the anti-PEG Ab ELISA assays. As for the inclusion criteria of blood donors, we had a mixed population “enriched” with sporadic and chronic allergic people and patients with mastocytosis.

The highly left-skewed distribution of pre-vaccination anti-PEG Abs in our study resembles the distribution of these Abs in patients treated with the PEGylated RNA aptamer, Pegnivacogin.^25^ On the other hand, our data may be the first to document such distribution in PEG-medication-free subjects. The finding of increased prevalence of anti-PEG Abs in females compared to males, a likely consequence of increased cosmetics use by women, is in line with many studies reporting such information.^21, 25, 26, 30-33^

### Anti-PEG IgG and IgM levels following vaccination with mRNA-LNP vaccines

Fig. 3A and B show the individual and median anti-PEG IgG and IgM levels after vaccination with mRNA vaccines as well as other, PEG-free Covid-19 vaccines (Sinopharm, Sputnik V and Astra Zeneca), used for controlling the effect of PEG. The pre-vaccination baseline, also shown in Fig. 1, displayed substantial individual variation for both Ab subclasses, relative to which highly significant rises after the first and 2 booster vaccinations were found only for Spikevax (Fig. 3A and B). For Comirnaty, we found significant post-vaccination increase only in anti-PEG IgM after the second jab (Fig. 3B). However, the considerable individual variation of anti-PEG Ab levels over a broad range both before and after vaccination, and the irrelevance of vaccination numbers when the question is the presence or absence of immunogenicity, led us to test another approach of analysis, relating the maximal postvaccination values to the pre-vaccination baselines for each vaccinees, regardless of injection number. This approach yielded highly significant rises of both anti-PEG IgG and IgM for Comirnaty, also (Fig 3C). As for other vaccines, the small number of preimmunization data did not yield statistically analyzable pre- and post-vaccination pairs.

**Figure 3.**
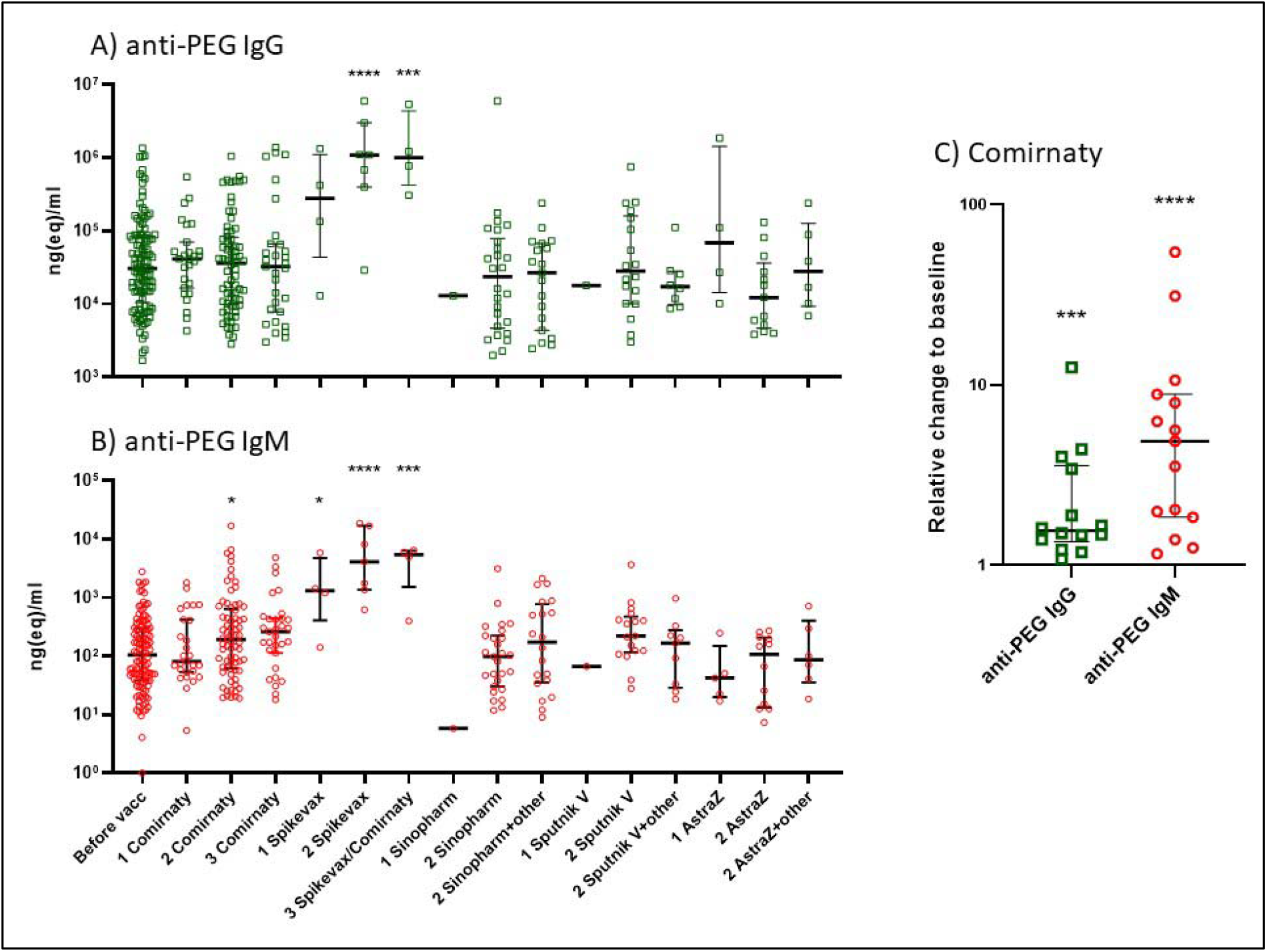
**Pre- and postvaccination anti-PEG Abs. Anti-PEG IgG (A, green rectangles) and anti-PEG IgM (B, red circles) levels in people vaccinated with different Covid-19 vaccines shown on the X axis, wherein the numbers (1, 2 and 3) represent the order of vaccinations. C) Relative rises of maximal postvaccination anti-PEG IgG and IgM levels relative to the respective preimmunization value in people undergoing serial (at least 2) immunizations with Comirnaty. Both absolute (A and B) and relative values (C) are presented on log scale. Of note, the time elapsed between the pre- and postvaccination measurements differ for each point. *, P< 0.05, ***, P< 0.001, ****, P<0.0001 by one-way ANOVA of logarithmic transformed data followed by Dunnett test (A and B) and Wilcoxon signed rank test of normalized data (C).**

Our finding that Spikevax induces highly significant rise of anti-PEG Abs is consistent with the mentioned recent data of Carreno et al.,^22^ and Ju et al,^21^ both groups coming to the same conclusion. Regarding our finding that Comirnaty, too, causes similar, although less expressed effect, which is seen only upon pairwise comparison, is also consistent with the latter two studies although both concluded that there was no such effect in case of Comirnaty.^21, 22^ However, a closer look at their data reveals very similar increases of anti-PEG IgG and IgM after vaccination with Comirnaty as we have seen in a pairwise comparison (Fig 3C). Notably, in the study of Carreno et al.^22^ Fig 2 shows 50-300% rise of prime/baseline anti-PEG IgG in 4/10 subjects, expressed as log AUC, and Fig S1 in the study of Ju et al.^21^ shows 13/17, 4/9 and 10/15 endpoint dilution value pairs in cohorts 1, 2 and 3, respectively, where the post-booster values visibly exceed the prevax values by > 20% up to several-fold on a log scale. These consistencies in experimental data from 3 research groups despite the fact all used different antigens in their ELISA (40 kDa-PEG,^21^ and 20kDa mPEG-BSA and 3.35-kDa free PEG^22^) provide strong indication for the occasional presence of a weak bystander anti-PEG immunogenicity of these vaccines. On the other end of the spectrum, the very high, outlier levels in a few anti-PEG Ab “supercarriers” can be rationalized with induced Abs whose extensive formation may involve positive feedback acceleration, a vicious cycle among anti-PEG Ab formation, C activation and HSRs.^41^

### Adverse reactions to Covid-19 mRNA-LNP vaccines

Table 2 categorizes all adverse events described by the 195 Comirnaty or Spikevax recipients of this study. Based on the definitions (see Table legend) we used for grouping the study participants according to reaction type, clinical grades and Brighton levels,^42^ 72% of vaccinees were reaction free (Grade 0), 20% gave account of usual vaccine side effects (Grade 1), and 8% reported Grade 2 or 3 HSR symptoms. There were more women among the reactors than man, and the 3 anaphylaxis cases were all women.

**Table 2.** Adverse reactions to mRNA/LNP Covid-19 vaccines.

| Clinically-based grading* | Symptom types | Brighton Levels** | Female | Male | All*** |
| --- | --- | --- | --- | --- | --- |
| Grade 0 | none | NA | 74 | 67 | 141 (72%) |
| Grade 1 | usual vaccine reactions |  | 28 | 11 | 39 (20%) |
| Grade 2 | hypersensitivity | 2-3 | 7 | 5 | 12 (6 %) |
| Grade 3 | reactions | 1 | 3 | 0 | 3 (2 %) |
| Total |  |  | 112 | 83 | 195 |

**\*Grade 0 is no adverse effect, Grade 1 "usual vaccine symptoms" include fever, arm pain, local redness, weakness, headache, chills, pruritus, depression, abdominal pain, indigestion, bloating, arthralgia, muscle pain, or fatigue. Grade 2 reactions represent allergic symptoms which pass spontaneously with or without epinephrine, antihistamines, or other pharmaceutical interventions. Grade 3 defines anaphylaxis or life-threatening reactions requiring emergency care (resuscitation and/or hospitalization). Grade 4, death, N/A in our study.**
**\*\*Level of diagnostic certainty for anaphylaxis, as defined in the Brighton Collaboration's anaphylaxis case definition guidelines.<sup>42</sup> According to this system level 1 "true anaphylaxis" is distinguished from other manifestations of allergy by its "diagnostic certainty", defined by a complex matrix of major and minor criteria. Our Grade 3 reactions roughly correspond to Brighton level 1, i.e., true anaphylaxis, wherein at least a major hemodynamic/circulatory or cardiopulmonary symptom (heart/back/limb pain, hypo- or hypertension, (angio)edema, swelling of the lips, tongue or face) and a major skin alteration (i.e., flushing, rash, erythema) are concurrently present. Brighton levels 2 and 3 (Grade 2) involve all other, non-life-threatening symptoms.**
**\*\*\*The % values represent prevalence in the 195 participants of this study involving both healthy and allergic people who were recalled for blood donation because of his/her HSR. Thus, the data do not represent classical epidemiological surveillance of vaccine adverse effects.**

### Hypersensitivity reactions to mRNA-LNP vaccines and their association with anti-PEG antibodies

The symptoms of 15 allergy responders included in this study are listed in Table 3, along with the gender, age range, the vaccine type and number, and the severity of HSRs.

**Table 3.**
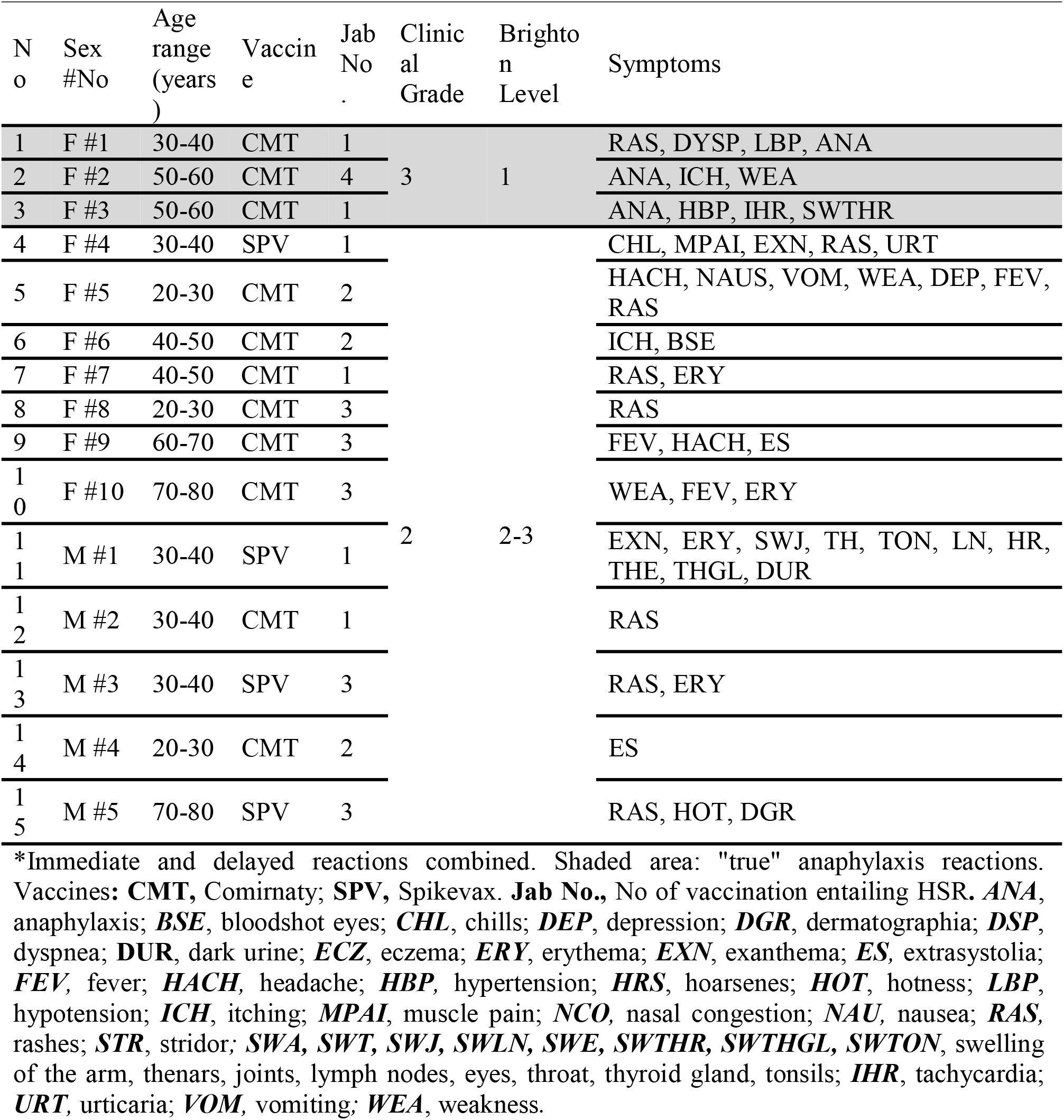
Hypersensitivity reactions in recipients of mRNA-containing Covid-19 vaccines*.

The above information in Table 3 reveals substantial individual variation of symptoms afflicting mainly the circulatory, cardio-pulmonary, the nerve systems and the skin. Figure 4 shows the anti-PEG IgG (A) and IgM (B) levels in reactor people displaying HSRs (R) compared to non-rector (NR) subjects. Both Ab levels were significantly higher in the R group compared to the NR, which suggests increased proneness of R people for anti-PEG Ab responses. However, because the blood withdrawals were not coincidental with the HSRs, temporal linkage between anti-PEG Ab rises and HSRs could not be assessed from these data.

**Figure 4.**
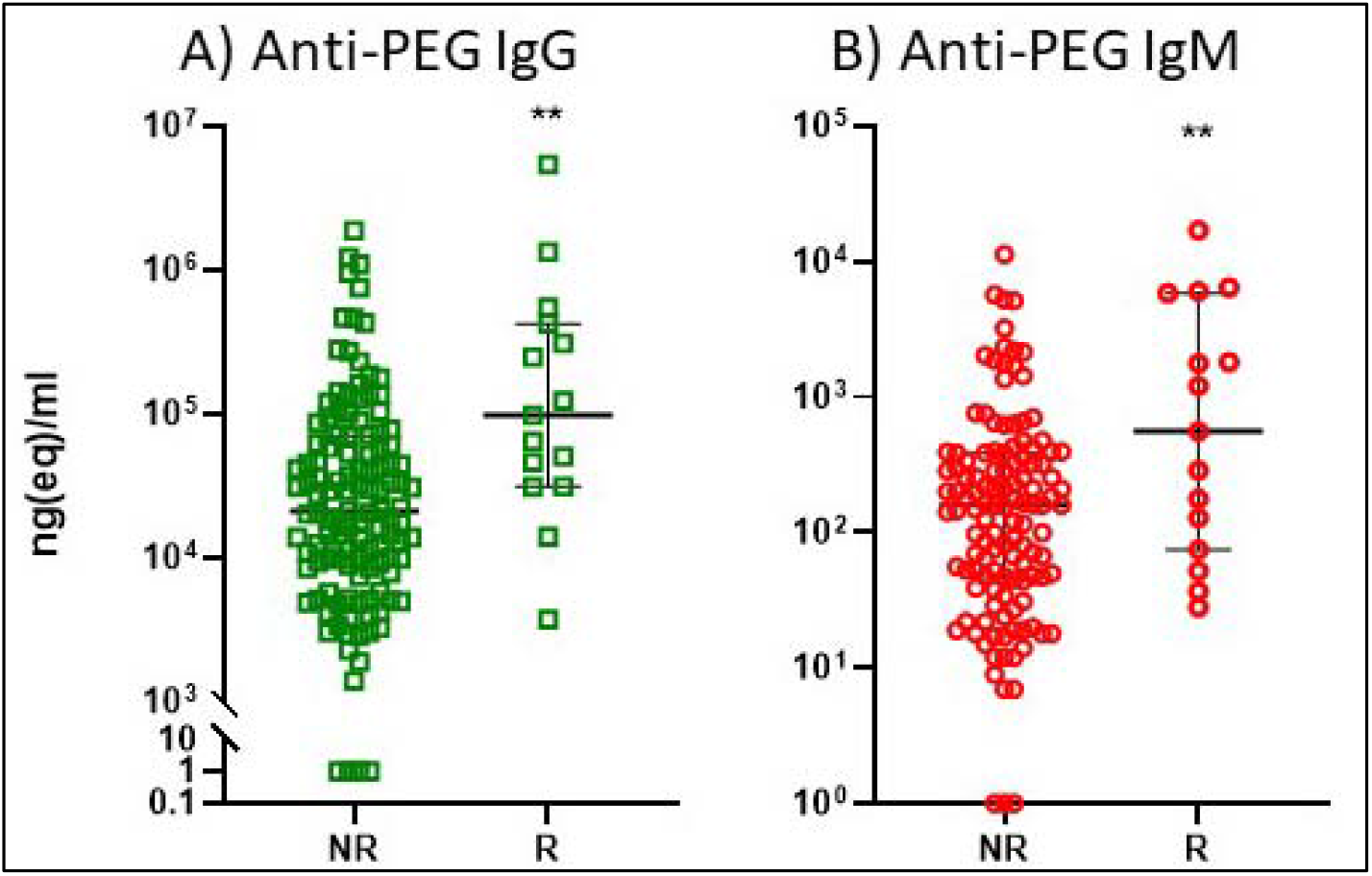
**Anti-PEG IgG (A), IgM (B) in reactor people displaying HSRs (R) compared to non-rector (NR) subjects. The symptoms observed in the NR (no adverse effects and Grade 1) and R (Grade 2 and 3) patients are detailed in Table 3. The time span between Ab determinations and vaccinations was 4 months. **P< 0.01 by t-Test of logarithmic transformed data.**

### SARS-CoV-2 neutralization antibody levels after multiple vaccinations: biphasic dependence on anti-PEG Ab levels

Serial measurements of anti-SARS-CoV-2 neutralizing, S protein-binding Abs (anti-S) in the plasma from 191 blood donors (n=361 tests) before and after vaccinations multiple times (in the 1-9 range) showed several versions of Ab responses, including small and large increases followed by steeper or weaker declines, constant low or high levels, or strong or weak initial or late responses (Supplement Table S1). This individual variation of antiviral immune response is in keeping with the variable and relatively short duration of immunity provided by the current vaccines, necessitation booster injections.

Plotting the anti-S levels versus the anti-PEG IgG and IgM after the first and second injections (Fig. 5A-D) in different subjects within 100 days after vaccination with Comirnaty (without any sign of Covid-19 infection) suggested bell-shaped curves, which could arise, among others, from a dose-dependent, biphasic correlation between anti-PEG Ab levels and anti-S immunogenicity. This phenomenon was best seen in the case of anti-PEG IgM after the second Comirnaty injection (Fig 5D).

**Figure 5.**
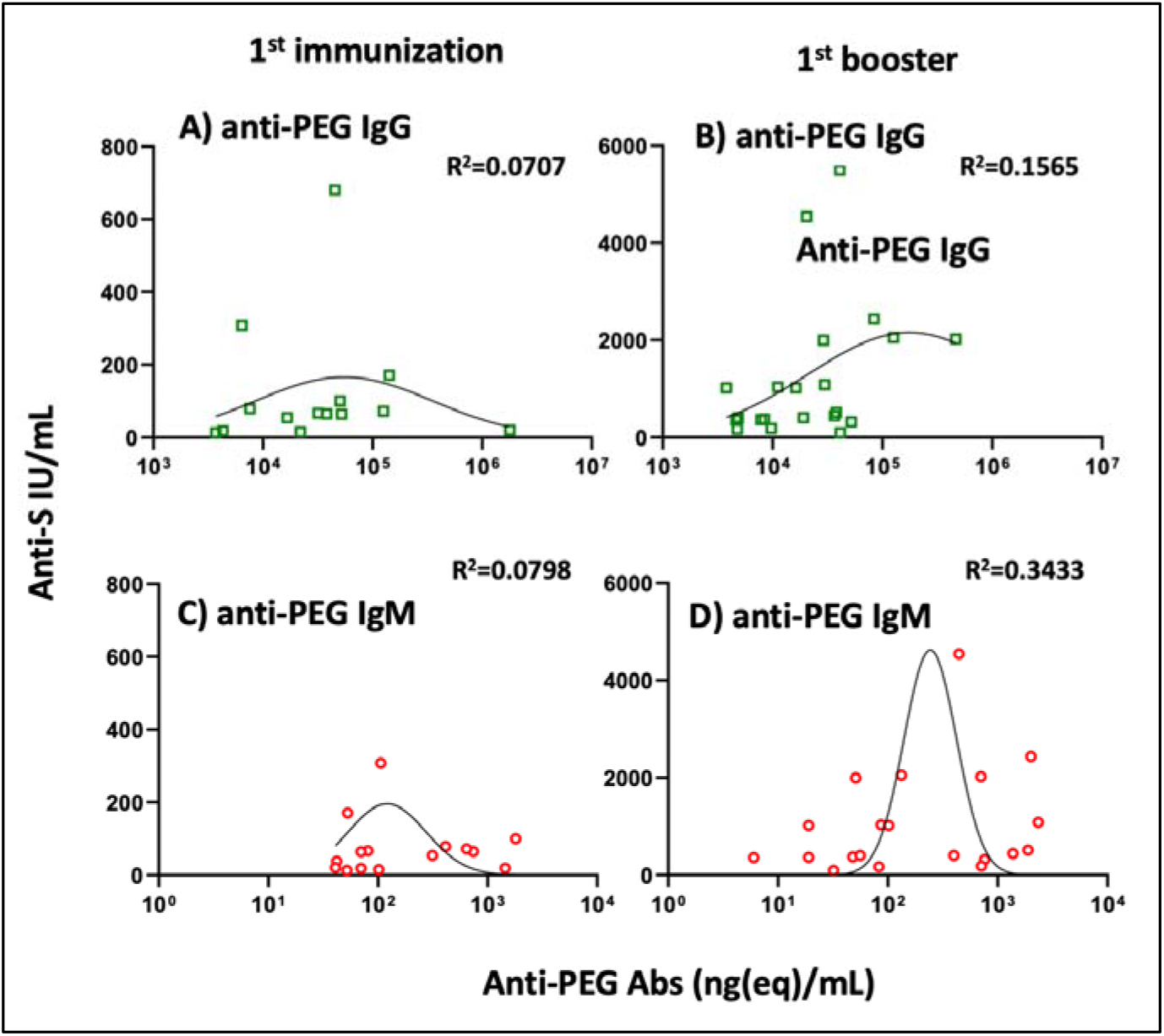
**Anti-S Ab levels as a function of anti-PEG Abs within 100 days after the first or the second vaccination with Comirnaty. Donors with any sign of Covid-19 infection were excluded. Each points represent the highest Anti-S value obtained in serial plasma samples in each individual. The goodness of lognormal Gaussian curve fitting is represented by R^2^ values on the figures.**

The paralleling increases in both Abs on the left-side of the bell curve may reflect increasing immune response against both PEG and the S-protein, while the waning levels of anti-S at higher anti-PEG levels may reflect interference by the binding of anti-PEG Abs to the vaccine NPs and possibly damage them. The latter assumption is based on the studies showing that the binding of anti-PEG Abs, particularly IgM, to PEGylated liposomes can cause complement activation with bilayer damage^41, 43^ as well as accelerated blood clearance of vesicles.^27-29^ Thus, anti-PEG Abs may represent an important biological variable which critically impacts the efficacy of mRNA-LNP vaccines in both directions.

## OUTLOOK

There is consensus in the literature that the incidence of HSRs and anaphylaxis caused by mRNA-LNP vaccinations is increased relative to that of traditional vaccines.^1-17^ Taken the ∼1.3 anaphylaxis/million out of >25 million recipients of flu vaccines,^44^ as reference, the extent of increase is in the 3-400-fold range (STable 2). Nevertheless, this potentially lethal side effect is still considered as very rare, and its occurrence is further decreasing with time, as precautions are increased.^18^ However, the >1 billion mRNA-LNP injections given worldwide^45^ places the sheer number of anaphylaxis cases probably in the tens of thousands (Table S2) and anaphylaxis is only the tip of the iceberg, a small section in a broad spectrum of HSR symptoms. These HSRs, even if mostly manageable, still represent a major acute health problem particularly for people with severe allergies, among whom the incidence of Comirnaty-indued anaphylaxis is 0.7% (STable 2), i.e., roughly 200-fold higher than in the normal population.^11^

We found in this study significantly increased levels of anti-PEG Abs in allergic reactor subjects compared to non-reactors, which finding, by itself, does not prove causal relationship between anti-PEG-Abs and HSRs. However, taken together with the unambiguous evidence for at least a contributing role of anti-PEG Abs to anaphylaxis, and also efficacy loss, in the discussed previous clinical and experiential studies with PEGylated NPs bearing similarities to mRNA-LNP,^23-29^ the warning by Ganson et al. “*we advise testing for pre-existing anti-PEG antibodies during clinical trials of new PEGylated therapeutic agents*”^25^ seems to be valid for mRNA-LNP vaccinations, too, particularly in light of the need for frequent booster injections. The potential anti-PEG bystander immunogenicity of these vaccines,^21, 22^ for which our study also provided evidence, represents another reason for attention to these Abs in the light of growing use of intrinsically reactogenic PEGylated nano-biopharmaceuticals. On the side of welcome news, it is likely that in the non-allergic population only anti-PEG Ab supercarrier people with very high levels of such Abs are at increased risk for HSRs and/or reduced efficacy of PEGylated vaccines or drugs.

## MATERIALS AND METHODS

### Materials

The Covid-19 vaccines administered to study participants and used in our in vitro studies were original, unexpired clinical batches. Their handling for storage and thawing was per the manufacturer’s recommendations. The ELISA kits for measuring SARS-CoV-2 neutralization antibody (TE 1076) were from TECO*Medical* AG, Sissach, Switzerland. Dulbecco’s phosphate-buffered saline (PBS) without Ca^++^/Mg^++^ and bovine calf serum, and biotin-labeled goat polyclonal anti-porcine IgM were from Sigma Chemical Co. (St. Louis, MO, USA).

### Procedures involving Humans

The volunteers for blood sample donations were between 8 months and 85 years of age and were vaccinated once or several times with combinations of mRNA (Comirnaty or Spikevax) and/or other vaccines, including Astra Zeneca, Jansen, Sputnik, and Sinopharm. A portion of subjects had immune disorders, i.e., allergies and mastocytosis. Blood was collected into EDTA vacutainers for plasma and clot activator tubes for serum. These were stored in aliquots at -80 °C until the various assays were performed. We also obtained samples from allergic reactors in other health care facilities, transported on dry ice. All participants, parents of minors, filled out a consent form and questionnaire asking about their age, medical and vaccination history, cosmetic use, and symptoms of HSRs, if they had experienced them after Covid-19 vaccination. The study was approved by the Scientific and Research Ethics Committee of the Hungarian Medical Research Council (52685-6/2022/EÜIG) and, for Miskolc University, BORS-02/2021.

### Antibody Tests

SARS-CoV-2 neutralization Abs specific against the receptor binding region of viral spike protein (anti-S) were measured as described in Refs. ^46, 47^ using a kit provided by TECO*medical* AG, (Sissach, Switzerland, *Catalog No. TE 1076*). Serial measurements of blood anti-PEG IgM and IgG levels were performed with ELISA as described earlier.^27, 36, 48^

### Statistical Analysis

The applied approaches, specified in the figure legends, included descriptive statistics, 2-tailed t-test on logarithmic transformed data, checking the normality of distribution by Shapiro-Wilk test, one-way ANOVA followed by Dunnett test, Wilcoxon signed rank test of normalized data and correlation analysis to test linear relationship between two variables.

## Data Availability

All data produced in the present study are available upon reasonable request to the authors

## ACKNOWLEDGMENTS

The authors are grateful to Marieluise Wippermann (TECO*Medical*, Sissach, Switzerland) for providing the SARS-CoV-2 neutralization kits. Thanks are due to Dr. Judit Varkonyi for recruiting patients with mastocytosis.

## FUNDING

The financial support by the European Union Horizon 2020 projects 825828 (Expert) and 952520 (Biosafety) are acknowledged. This project was supported by a grant from the National Research, Development, and Innovation Office (NKFIH) of Hungary (2020-1.1.6-JÖVŐ-2021-00013). JS thanks the logistic support by the Applied Materials and Nanotechnology, Center of Excellence, Miskolc University, Miskolc, Hungary.

## CONFLICT OF INTEREST

The authors affiliated with SeroScience LLC are involved in the company’s contract research service activity providing studies that were applied here. The funders had no role in the design of the study; in the collection, analyses, or interpretation of data; in the writing of the manuscript, or in the decision to publish the results.

## SUPPLEMENTARY INFORMATION

**Figure S1.**
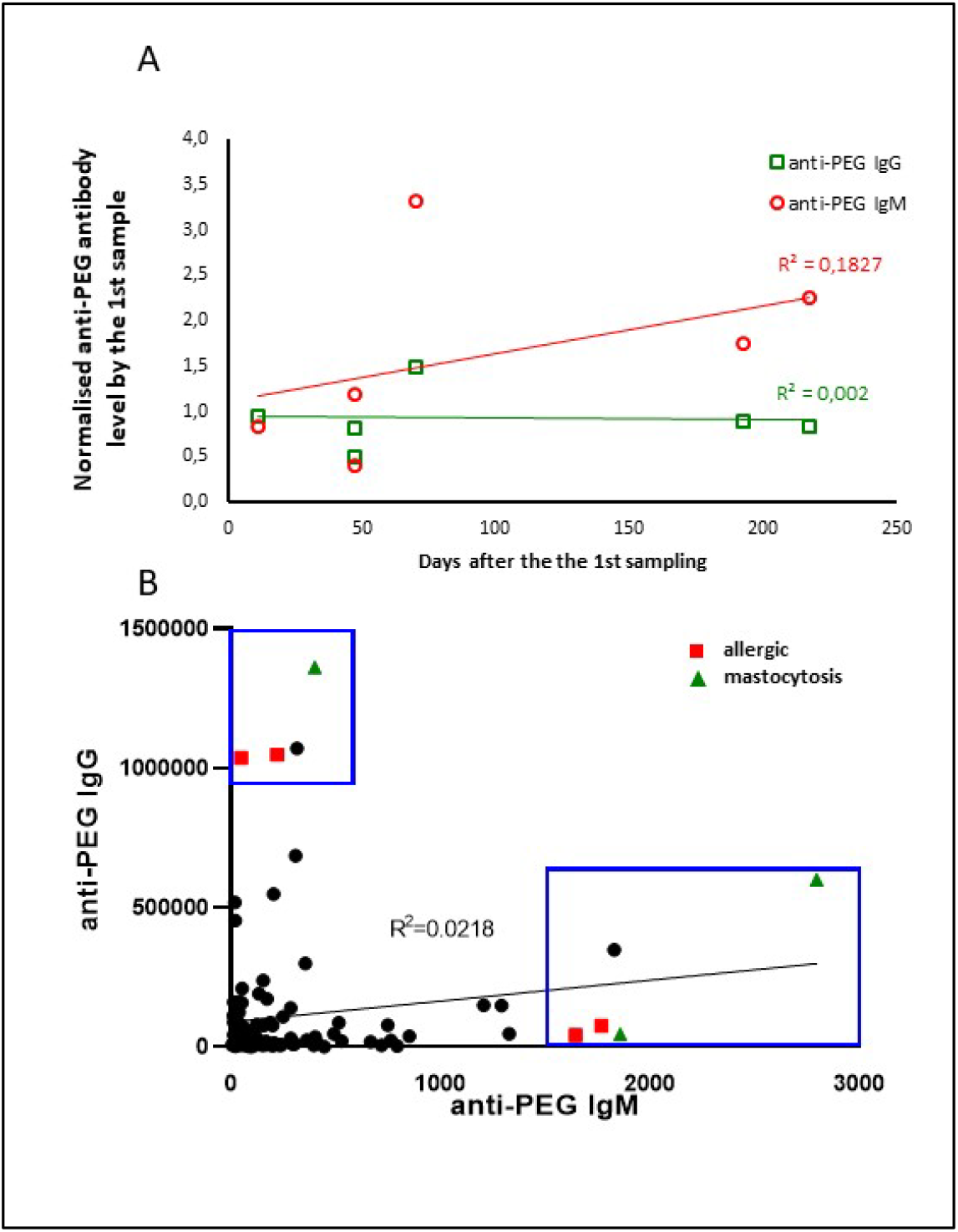
**Anti-PEG Ab levels in peopl,e before Covid-19 vaccinations. A) Changes over time in 6 subjects who gave blood 2 times before vaccination against Covid-19. The second/first anti-PEG IgG and IgM ratios were plotted against the time elapsed time between the 1st and 2nd blood withdrawal. B) Lack of correlation of plasma anti-PEG-IgM and anti-PEG-IgG in all blood donors before vacination (black dots), except in those with allergy (red squares) or mastocytosis (green triangles), in whom maximal levels of anti-PEG IgG were associated with minimal values of anti-PEG IgM. Antibody levels were determined as described in the Methods.**

**Table S1.**
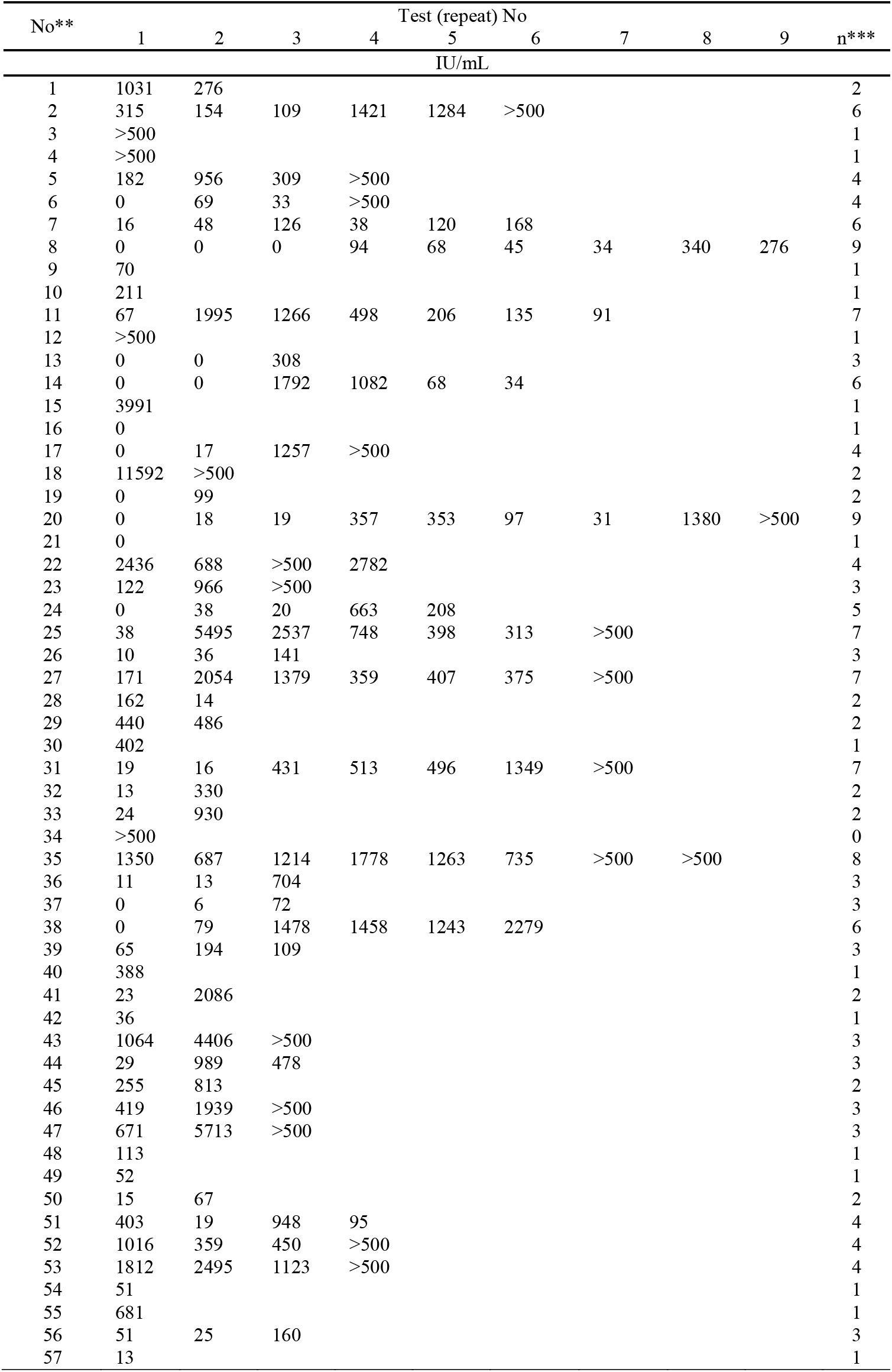
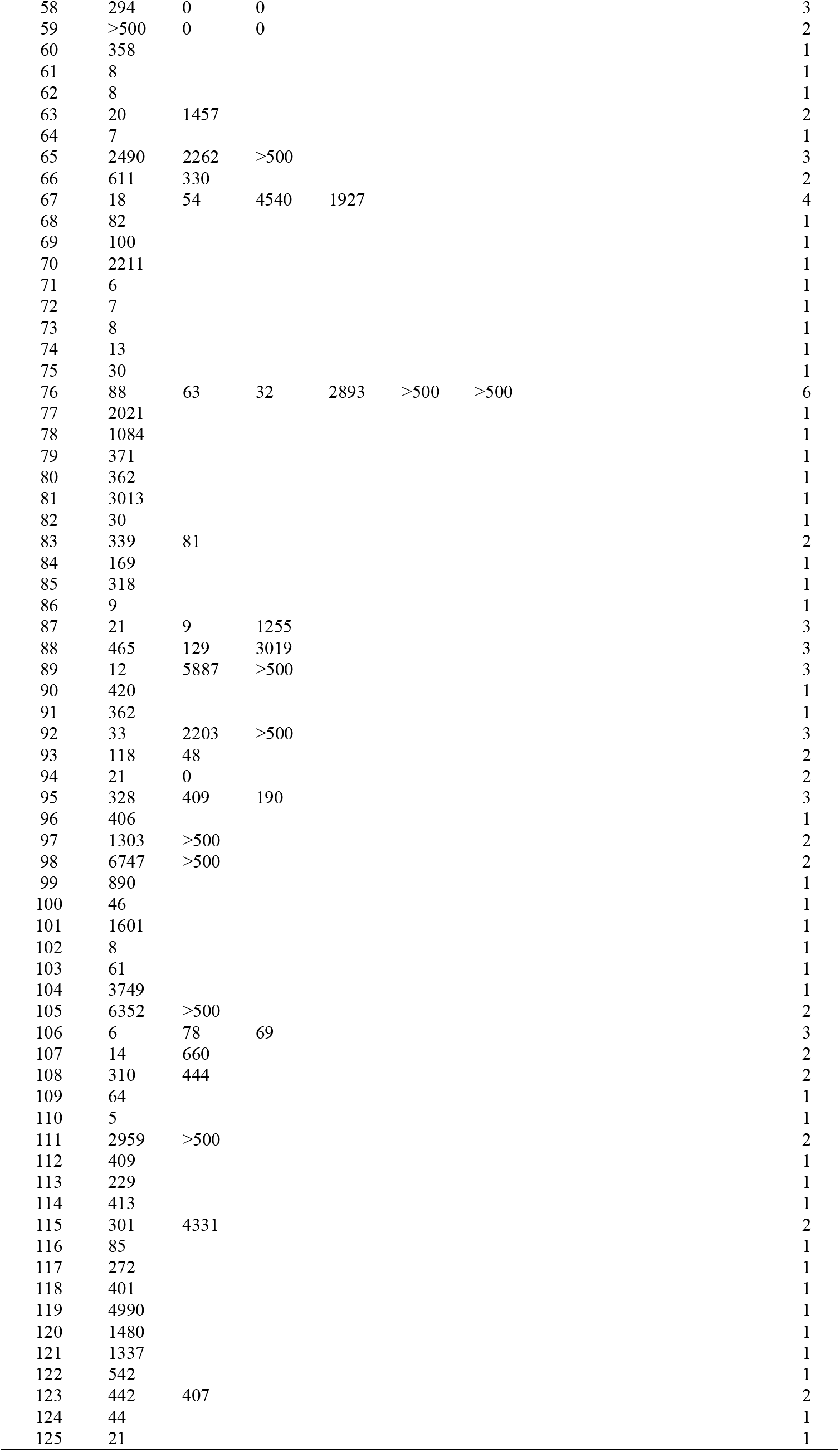
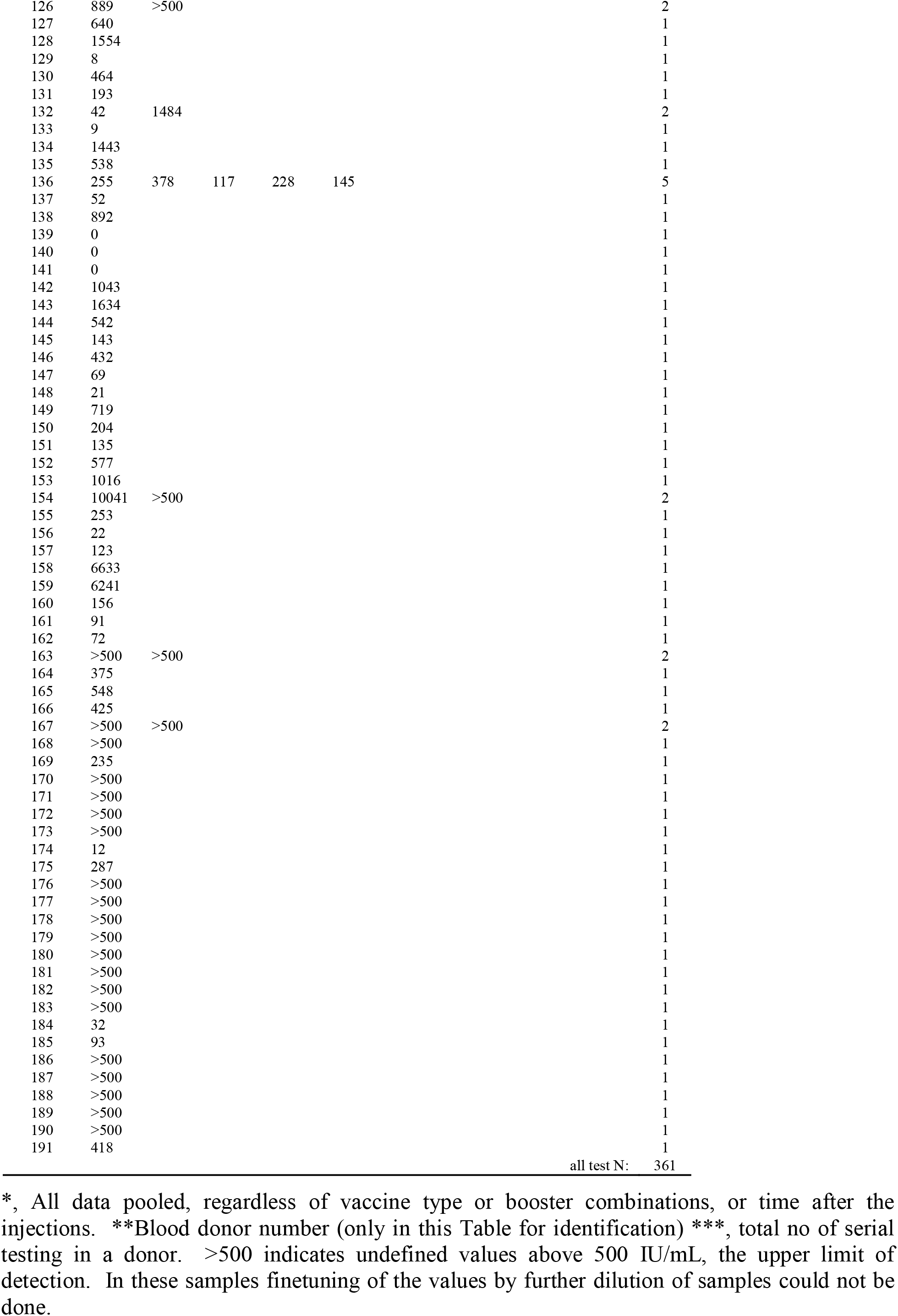
Inter- and intra-individual variation of anti-S levels in Covid-19 vaccine recipients*.

| No** | Test (repeat) No |  |  |  |  |  |  |  |  | n*** |
| --- | --- | --- | --- | --- | --- | --- | --- | --- | --- | --- |
|  | 1 | 2 | 3 | 4 | 5 | 6 | 7 | 8 | 9 |  |
|  | IU/mL |  |  |  |  |  |  |  |  |  |
| 1 | 1031 | 276 |  |  |  |  |  |  |  | 2 |
| 2 | 315 | 154 | 109 | 1421 | 1284 | >500 |  |  |  | 6 |
| 3 | >500 |  |  |  |  |  |  |  |  | 1 |
| 4 | >500 |  |  |  |  |  |  |  |  | 1 |
| 5 | 182 | 956 | 309 | >500 |  |  |  |  |  | 4 |
| 6 | 0 | 69 | 33 | >500 |  |  |  |  |  | 4 |
| 7 | 16 | 48 | 126 | 38 | 120 | 168 |  |  |  | 6 |
| 8 | 0 | 0 | 0 | 94 | 68 | 45 | 34 | 340 | 276 | 9 |
| 9 | 70 |  |  |  |  |  |  |  |  | 1 |
| 10 | 211 |  |  |  |  |  |  |  |  | 1 |
| 11 | 67 | 1995 | 1266 | 498 | 206 | 135 | 91 |  |  | 7 |
| 12 | >500 |  |  |  |  |  |  |  |  | 1 |
| 13 | 0 | 0 | 308 |  |  |  |  |  |  | 3 |
| 14 | 0 | 0 | 1792 | 1082 | 68 | 34 |  |  |  | 6 |
| 15 | 3991 |  |  |  |  |  |  |  |  | 1 |
| 16 | 0 |  |  |  |  |  |  |  |  | 1 |
| 17 | 0 | 17 | 1257 | >500 |  |  |  |  |  | 4 |
| 18 | 11592 | >500 |  |  |  |  |  |  |  | 2 |
| 19 | 0 | 99 |  |  |  |  |  |  |  | 2 |
| 20 | 0 | 18 | 19 | 357 | 353 | 97 | 31 | 1380 | >500 | 9 |
| 21 | 0 |  |  |  |  |  |  |  |  | 1 |
| 22 | 2436 | 688 | >500 | 2782 |  |  |  |  |  | 4 |
| 23 | 122 | 966 | >500 |  |  |  |  |  |  | 3 |
| 24 | 0 | 38 | 20 | 663 | 208 |  |  |  |  | 5 |
| 25 | 38 | 5495 | 2537 | 748 | 398 | 313 | >500 |  |  | 7 |
| 26 | 10 | 36 | 141 |  |  |  |  |  |  | 3 |
| 27 | 171 | 2054 | 1379 | 359 | 407 | 375 | >500 |  |  | 7 |
| 28 | 162 | 14 |  |  |  |  |  |  |  | 2 |
| 29 | 440 | 486 |  |  |  |  |  |  |  | 2 |
| 30 | 402 |  |  |  |  |  |  |  |  | 1 |
| 31 | 19 | 16 | 431 | 513 | 496 | 1349 | >500 |  |  | 7 |
| 32 | 13 | 330 |  |  |  |  |  |  |  | 2 |
| 33 | 24 | 930 |  |  |  |  |  |  |  | 2 |
| 34 | >500 |  |  |  |  |  |  |  |  | 0 |
| 35 | 1350 | 687 | 1214 | 1778 | 1263 | 735 | >500 | >500 |  | 8 |
| 36 | 11 | 13 | 704 |  |  |  |  |  |  | 3 |
| 37 | 0 | 6 | 72 |  |  |  |  |  |  | 3 |
| 38 | 0 | 79 | 1478 | 1458 | 1243 | 2279 |  |  |  | 6 |
| 39 | 65 | 194 | 109 |  |  |  |  |  |  | 3 |
| 40 | 388 |  |  |  |  |  |  |  |  | 1 |
| 41 | 23 | 2086 |  |  |  |  |  |  |  | 2 |
| 42 | 36 |  |  |  |  |  |  |  |  | 1 |
| 43 | 1064 | 4406 | >500 |  |  |  |  |  |  | 3 |
| 44 | 29 | 989 | 478 |  |  |  |  |  |  | 3 |
| 45 | 255 | 813 |  |  |  |  |  |  |  | 2 |
| 46 | 419 | 1939 | >500 |  |  |  |  |  |  | 3 |
| 47 | 671 | 5713 | >500 |  |  |  |  |  |  | 3 |
| 48 | 113 |  |  |  |  |  |  |  |  | 1 |
| 49 | 52 |  |  |  |  |  |  |  |  | 1 |
| 50 | 15 | 67 |  |  |  |  |  |  |  | 2 |
| 51 | 403 | 19 | 948 | 95 |  |  |  |  |  | 4 |
| 52 | 1016 | 359 | 450 | >500 |  |  |  |  |  | 4 |
| 53 | 1812 | 2495 | 1123 | >500 |  |  |  |  |  | 4 |
| 54 | 51 |  |  |  |  |  |  |  |  | 1 |
| 55 | 681 |  |  |  |  |  |  |  |  | 1 |
| 56 | 51 | 25 | 160 |  |  |  |  |  |  | 3 |
| 57 | 13 |  |  |  |  |  |  |  |  | 1 |

|  |  |  |  |  |  |  |  |
| --- | --- | --- | --- | --- | --- | --- | --- |
| 58 | 294 | 0 | 0 |  |  |  | 3 |
| 59 | >500 | 0 | 0 |  |  |  | 2 |
| 60 | 358 |  |  |  |  |  | 1 |
| 61 | 8 |  |  |  |  |  | 1 |
| 62 | 8 |  |  |  |  |  | 1 |
| 63 | 20 | 1457 |  |  |  |  | 2 |
| 64 | 7 |  |  |  |  |  | 1 |
| 65 | 2490 | 2262 | >500 |  |  |  | 3 |
| 66 | 611 | 330 |  |  |  |  | 2 |
| 67 | 18 | 54 | 4540 | 1927 |  |  | 4 |
| 68 | 82 |  |  |  |  |  | 1 |
| 69 | 100 |  |  |  |  |  | 1 |
| 70 | 2211 |  |  |  |  |  | 1 |
| 71 | 6 |  |  |  |  |  | 1 |
| 72 | 7 |  |  |  |  |  | 1 |
| 73 | 8 |  |  |  |  |  | 1 |
| 74 | 13 |  |  |  |  |  | 1 |
| 75 | 30 |  |  |  |  |  | 1 |
| 76 | 88 | 63 | 32 | 2893 | >500 | >500 | 6 |
| 77 | 2021 |  |  |  |  |  | 1 |
| 78 | 1084 |  |  |  |  |  | 1 |
| 79 | 371 |  |  |  |  |  | 1 |
| 80 | 362 |  |  |  |  |  | 1 |
| 81 | 3013 |  |  |  |  |  | 1 |
| 82 | 30 |  |  |  |  |  | 1 |
| 83 | 339 | 81 |  |  |  |  | 2 |
| 84 | 169 |  |  |  |  |  | 1 |
| 85 | 318 |  |  |  |  |  | 1 |
| 86 | 9 |  |  |  |  |  | 1 |
| 87 | 21 | 9 | 1255 |  |  |  | 3 |
| 88 | 465 | 129 | 3019 |  |  |  | 3 |
| 89 | 12 | 5887 | >500 |  |  |  | 3 |
| 90 | 420 |  |  |  |  |  | 1 |
| 91 | 362 |  |  |  |  |  | 1 |
| 92 | 33 | 2203 | >500 |  |  |  | 3 |
| 93 | 118 | 48 |  |  |  |  | 2 |
| 94 | 21 | 0 |  |  |  |  | 2 |
| 95 | 328 | 409 | 190 |  |  |  | 3 |
| 96 | 406 |  |  |  |  |  | 1 |
| 97 | 1303 | >500 |  |  |  |  | 2 |
| 98 | 6747 | >500 |  |  |  |  | 2 |
| 99 | 890 |  |  |  |  |  | 1 |
| 100 | 46 |  |  |  |  |  | 1 |
| 101 | 1601 |  |  |  |  |  | 1 |
| 102 | 8 |  |  |  |  |  | 1 |
| 103 | 61 |  |  |  |  |  | 1 |
| 104 | 3749 |  |  |  |  |  | 1 |
| 105 | 6352 | >500 |  |  |  |  | 2 |
| 106 | 6 | 78 | 69 |  |  |  | 3 |
| 107 | 14 | 660 |  |  |  |  | 2 |
| 108 | 310 | 444 |  |  |  |  | 2 |
| 109 | 64 |  |  |  |  |  | 1 |
| 110 | 5 |  |  |  |  |  | 1 |
| 111 | 2959 | >500 |  |  |  |  | 2 |
| 112 | 409 |  |  |  |  |  | 1 |
| 113 | 229 |  |  |  |  |  | 1 |
| 114 | 413 |  |  |  |  |  | 1 |
| 115 | 301 | 4331 |  |  |  |  | 2 |
| 116 | 85 |  |  |  |  |  | 1 |
| 117 | 272 |  |  |  |  |  | 1 |
| 118 | 401 |  |  |  |  |  | 1 |
| 119 | 4990 |  |  |  |  |  | 1 |
| 120 | 1480 |  |  |  |  |  | 1 |
| 121 | 1337 |  |  |  |  |  | 1 |
| 122 | 542 |  |  |  |  |  | 1 |
| 123 | 442 | 407 |  |  |  |  | 2 |
| 124 | 44 |  |  |  |  |  | 1 |
| 125 | 21 |  |  |  |  |  | 1 |
| 126 | 889 | >500 |  |  |  |  | 2 |
| 127 | 640 |  |  |  |  |  | 1 |
| 128 | 1554 |  |  |  |  |  | 1 |
| 129 | 8 |  |  |  |  |  | 1 |
| 130 | 464 |  |  |  |  |  | 1 |
| 131 | 193 |  |  |  |  |  | 1 |
| 132 | 42 | 1484 |  |  |  |  | 2 |
| 133 | 9 |  |  |  |  |  | 1 |
| 134 | 1443 |  |  |  |  |  | 1 |
| 135 | 538 |  |  |  |  |  | 1 |
| 136 | 255 | 378 | 117 | 228 | 145 |  | 5 |
| 137 | 52 |  |  |  |  |  | 1 |
| 138 | 892 |  |  |  |  |  | 1 |
| 139 | 0 |  |  |  |  |  | 1 |
| 140 | 0 |  |  |  |  |  | 1 |
| 141 | 0 |  |  |  |  |  | 1 |
| 142 | 1043 |  |  |  |  |  | 1 |
| 143 | 1634 |  |  |  |  |  | 1 |
| 144 | 542 |  |  |  |  |  | 1 |
| 145 | 143 |  |  |  |  |  | 1 |
| 146 | 432 |  |  |  |  |  | 1 |
| 147 | 69 |  |  |  |  |  | 1 |
| 148 | 21 |  |  |  |  |  | 1 |
| 149 | 719 |  |  |  |  |  | 1 |
| 150 | 204 |  |  |  |  |  | 1 |
| 151 | 135 |  |  |  |  |  | 1 |
| 152 | 577 |  |  |  |  |  | 1 |
| 153 | 1016 |  |  |  |  |  | 1 |
| 154 | 10041 | >500 |  |  |  |  | 2 |
| 155 | 253 |  |  |  |  |  | 1 |
| 156 | 22 |  |  |  |  |  | 1 |
| 157 | 123 |  |  |  |  |  | 1 |
| 158 | 6633 |  |  |  |  |  | 1 |
| 159 | 6241 |  |  |  |  |  | 1 |
| 160 | 156 |  |  |  |  |  | 1 |
| 161 | 91 |  |  |  |  |  | 1 |
| 162 | 72 |  |  |  |  |  | 1 |
| 163 | >500 | >500 |  |  |  |  | 2 |
| 164 | 375 |  |  |  |  |  | 1 |
| 165 | 548 |  |  |  |  |  | 1 |
| 166 | 425 |  |  |  |  |  | 1 |
| 167 | >500 | >500 |  |  |  |  | 2 |
| 168 | >500 |  |  |  |  |  | 1 |
| 169 | 235 |  |  |  |  |  | 1 |
| 170 | >500 |  |  |  |  |  | 1 |
| 171 | >500 |  |  |  |  |  | 1 |
| 172 | >500 |  |  |  |  |  | 1 |
| 173 | >500 |  |  |  |  |  | 1 |
| 174 | 12 |  |  |  |  |  | 1 |
| 175 | 287 |  |  |  |  |  | 1 |
| 176 | >500 |  |  |  |  |  | 1 |
| 177 | >500 |  |  |  |  |  | 1 |
| 178 | >500 |  |  |  |  |  | 1 |
| 179 | >500 |  |  |  |  |  | 1 |
| 180 | >500 |  |  |  |  |  | 1 |
| 181 | >500 |  |  |  |  |  | 1 |
| 182 | >500 |  |  |  |  |  | 1 |
| 183 | >500 |  |  |  |  |  | 1 |
| 184 | 32 |  |  |  |  |  | 1 |
| 185 | 93 |  |  |  |  |  | 1 |
| 186 | >500 |  |  |  |  |  | 1 |
| 187 | >500 |  |  |  |  |  | 1 |
| 188 | >500 |  |  |  |  |  | 1 |
| 189 | >500 |  |  |  |  |  | 1 |
| 190 | >500 |  |  |  |  |  | 1 |
| 191 | 418 |  |  |  |  |  | 1 |

\*, All data pooled, regardless of vaccine type or booster combinations, or time after the injections. \*\*Blood donor number (only in this Table for identification) \*\*\*, total no of serial testing in a donor. >500 indicates undefined values above 500 IU/mL, the upper limit of detection. In these samples finetuning of the values by further dilution of samples could not be done.

**Table S2.** Different statistics on the incidence of anaphylaxis to mRNA-LNP vaccines.

| **Vaccine** | **Reaction n\*** | **Total n\*\*** | **Ana/million** | **Ana %** | **References** |
| --- | --- | --- | --- | --- | --- |
| Comirnaty | 47 | 9,943,247 | 5 | 0.00047 | 5, 49 |
| Spikevax | 19 | 7,581,429 | 3 | 0.00025 | 4, 50 |
| Comirnaty | 46 | 14,475,979 | 3 | 0.00032 | 18 |
| Comirnaty | 3730 | 242,000,000 | 15 | 0.00154 | 48 |
| Spikevax | 1455 | 154,000,000 | 9 | 0.00094 |  |
| Comirnaty + Spikevax | 1 | 4000 | 250 | 0.02500 | 8 |
| Comirnaty | 7 | 25929 | 270 | 0.02700 | 9 |
| Spikevax | 9 | 38971 | 231 | 0.02309 |  |
| Comirnaty | 14 | 31635 | 443 | 0.04425 | 3 |
| Spikevax | 3 | 7260 | 413 | 0.04132 |  |
| Comirnaty | 3 | 429*** | 2,331 | 0.699 | 11 |
|  |  | 80,943**** | 37 | 0.00371 |  |

\*, Number of reported anaphylaxis or "severe" HSR cases; \*\*, Total n of injections or vaccine recipients; \*\*\*, Only highly allergic people were included in the statistic; \*\*\*\*, Normalized to the healthy population based on the ratio of highly allergic people among people with any allergic complain (5,3%)<sup>11</sup> and a rough estimate of 10% prevalence of different allergies worldwide (Wikipedia).

## Notes

### Competing Interest Statement

The authors have declared no competing interest.

### Funding Statement

This study was funded by EU Horizon 2020

### Author Declarations

Ethics Committee of Miskolc University gave ethical approval for this work

